# A playbook for harmonization of standards for emergent viral pathogens

**DOI:** 10.1101/2024.10.05.24314949

**Authors:** Sara Suliman, Sebastien Fuchs, David Catoe, Loren Hansen, Deepa Eveleigh, Michael A. Crone, Paul S. Freemont, Martin Kammel, Heinz Zeichhardt, Andrew T. Anfora, Russell Garlick, Neil Almond, Mark Page, Brian Beck, Hui Wang, Karuna Sharma, Gary Pestano, Joannes Vancann, Jo Vandesompele, Adam S Corner, Jeffrey Albrecht, Deborah J. Boles, Marcia Eisenberg, Ayla B. Harris, Brian J. Krueger, Amanda L. Suchanek, Thomas J. Urban, Jonathan D. Williams, Patrick Chain, Alina Deshpande, Attelia Hollander, Sujata Chalise, David R. Walt, Jeffrey Germer, Joseph D. Yao, Bobbi S. Pritt, Frederick S. Nolte, Alexandra Bogožalec Košir, Mojca Milavec, Megan H. Cleveland, Peter M. Vallone, Eloise J. Busby, Jim F. Huggett, Denise M. O’Sullivan, Elizabeth M. Marlowe, Benjamin A. Pinsky, Annaleise Kealiher, Jeantine E. Lunshof, Christopher E. Mason, John Sninsky, Cristina Tato, Timothy Mercer, Thomas J. White, Marc Salit

## Abstract

Effective clinical and public health decision-making during a pandemic depends on reliable and interoperable clinical diagnostic test results. To ensure trustworthy outcomes, we need widely available and harmonized calibration standards on a shared scale. We present a ‘playbook’ for an inter-laboratory harmonization study that calibrates any available standards against a limited-availability standard issued by a global authority like the World Health Organization (WHO).

## Main Text

### What this is

As part of the global response to the COVID-19 pandemic^1,2^, various organizations rapidly made available a wide variety of standards and controls to develop, calibrate, and validate molecular diagnostics for SARS-CoV-2. Our *Coronavirus* Standards *Working Group* (CSWG)^3^ designed and conducted a “harmonization study” to establish the comparability of a set of these standards and controls (hereafter collectively referred to as ‘standards’). Harmonized standards can be used as a basis for meaningful quantitative comparability of test performance, and can underpin quantitative virology, therapeutic actions, and vaccinology. Harmonization balances the tension between the widespread need for standards and the limited availability of standards from global authorities. Our study is presented here in the form of a “playbook” as a recipe to conduct a robust harmonization study when it is needed again.

We calibrated 8 widely available molecular standards to the rapidly developed World Health Organization (WHO) International Standard (IS)^4^, putting all the quantitative values of these standards on this common scale **(Supplementary Table 1A)**. We recruited 14 laboratories internationally to participate in a designed experiment to “value assign” the standards on the scale **(Figure 1A and B and Supplementary Table 1B)**. Our intent was for widely available harmonized standards to be part of the infrastructure to confidently deploy testing at scale. We prototyped a “proof-of-concept study” to test whether calibrating with the harmonized standards would yield comparable results when measuring clinical samples **(Supplementary Figure 1)**.

**Figure 1:**
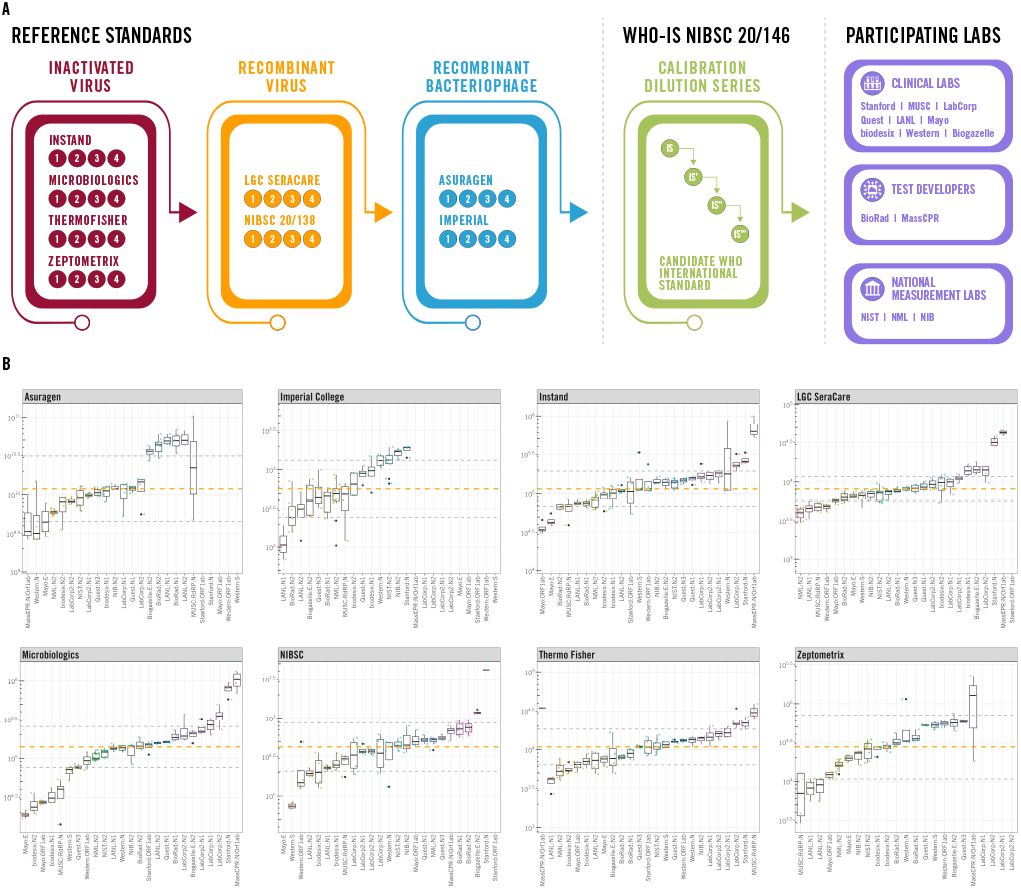
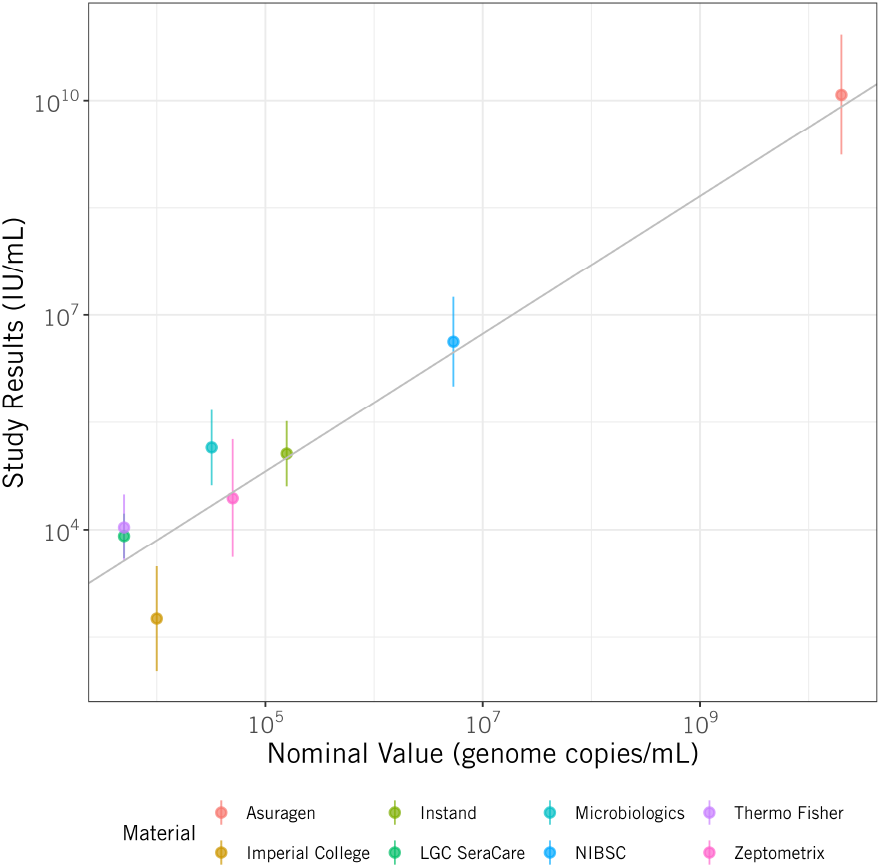
**(A)** Molecular Standards Harmonization Study Design: Schematic representation of the Coronavirus Standards Working Group (CSWG) molecular standards harmonization study. Four identical aliquots of each of the secondary reference materials were shipped to the 14 participating laboratories. The NIBSC code 20/146 WHO-IS was also shipped in 4 aliquots to benchmark the 8 secondary reference standards. **(B)** Box and whiskers represent median and interquartile ranges corresponding to the SARS-CoV-2 concentrations in IU, calibrated using the NIBSC 20/146 reference material. Each measurement represents a unique laboratory and assay, with the orange line spanning the median concentration across the different platforms and assays. Measurements are ordered from the lowest to highest extrapolated values. Data are summarized assuming each laboratory:assay are independent samples from the population. **(C)** Observed concentrations of each standard in log_10_ IU mL^-1^ versus the nominal value in log_10_ genome copies mL^-1^. The error bars are 95% Confidence Intervals. The line represents the IS values for the international standard serial dilution. The line is the multiplicative factor of the IS value to copies, showing that this proportionality is consistent across the population of the 8 standards.

This interlaboratory study had the sole aim to establish the quantitative comparability of these widely available SARS-CoV-2 standards for molecular diagnostics. The study was not intended to be a comparison of tests or laboratories, a survey of test performance, or an evaluation of commutability.

### Why it matters

The pandemic response (including development of diagnostics) was based on best practices developed in the moment. We offer here a study design pre-positioned for a better response to emerging pathogens with pandemic potential. Harmonizing standards with this design will assure comparability of results from diagnostic tests^5^. Comparable results enable quantitative, systematic diagnostic virology to understand pathogenesis and rapidly integrate data across space and time. Understanding pathogenesis^6^ and transmission^2,7-9^ is critical for clinical and public health decision-making, and well-calibrated controls are essential for adjudicating accuracy of diagnostics, especially tests using new or nascent technology. We present and support this approach here for a viral pathogen but recognize that the principles would apply to non-viral pathogens.

### What we did

The study was coordinated by the Joint Initiative for Metrology in Biology, the convener of the CSWG^3^. The coordinating laboratory worked with the CSWG to design the study, develop a Standard Operating Procedure (SOP), and create a reporting template for the study. The study design ensured that the results meaningfully represented the variety of standard materials and laboratories and their protocols **(Figure 1A)**. The SOP ensured that we obtained results from a common experimental design that permitted assessment of variability **(Supplement 1)**, and the template ensured consistent analysis of results **(Supplementary Table 2 and Methods)**. We developed an open public web-based analysis package to transparently present and share results (Shiny App: https://msalit.shinyapps.io/RNAstudy/).

We selected 8 widely available standards (including materials derived from inactivated virus, recombinant virus, and recombinant bacteriophage) that could be processed in the laboratory workflow as clinical specimens. We recruited 14 laboratories from different sectors (clinical, academic, public, commercial, and government) to each calibrate the standards to Medicines and Healthcare Products Regulatory Agency (MHRA), formerly known as National Institute for Biological Standards and Control (NIBSC) 20/146^4^, the first WHO International Standard for SARS-CoV-2 RNA (WHO-IS) that defined the International Unit (IU, **Figure 1C, Supplementary Table 1A**). Our study design used 4 replicate serial dilutions of the IS across 6 orders of magnitude, with 4 replicates of each dilution of the WHO-IS and each standard **(Supplementary Figures 2A-K)**. After calibration of these 8 standards, they were harmonized to a common scale by their value assignment in IU.

We operated a companion study to evaluate the hypothesis that the PCR tests calibrated with harmonized standards yield comparable results when measuring clinical samples **(Supplementary Figure 1)**. We used pooled biobanked samples (15 pools including negative controls and clinical positives at 3 viral load levels), evaluated by a subset of laboratories (3 laboratories using a total of 5 different assays) and two of the 8 standards.

### How it came out

#### Harmonization results

The 8 standards were all successfully value-assigned to a common, shared reference: the IU (**Figure 1C** and *Results v. Nominal* in analysis dashboard of the Shiny app). The analysis dashboard also contains figures showing the measurement results for each standard by assay within lab for each of the 8 materials sortable by rank order or laboratory; deviations from median value for each assay; tabular results, and raw data.

#### Proof-of-Concept results

Calibrating the assays with harmonized standards allowed us to compare results from different laboratories and tests on the clinical samples **(Figure 2A)**. Each laboratory used its own experiment design, representing real-life testing where labs use different tests. This proof-of-concept experiment demonstrated the use of calibration to bring clinical sample results from disparate assay modalities (reverse transcription quantitative real time PCR (RT-qPCR) and reverse transcription digital PCR (RT-dPCR)) to the same scale, plotted on the same axis **(Figure 2B)**. Critical analysis of these data and assessment of bias and variability is in the Supplement **(Supplementary Figure 3A-C)**. Lessons learned from this experiment have been incorporated into the playbook recommendations. Results from these studies promise that test results can be aggregated across space-and-time for scientific knowledge development and public-health decision-making.

**Figure 2:**
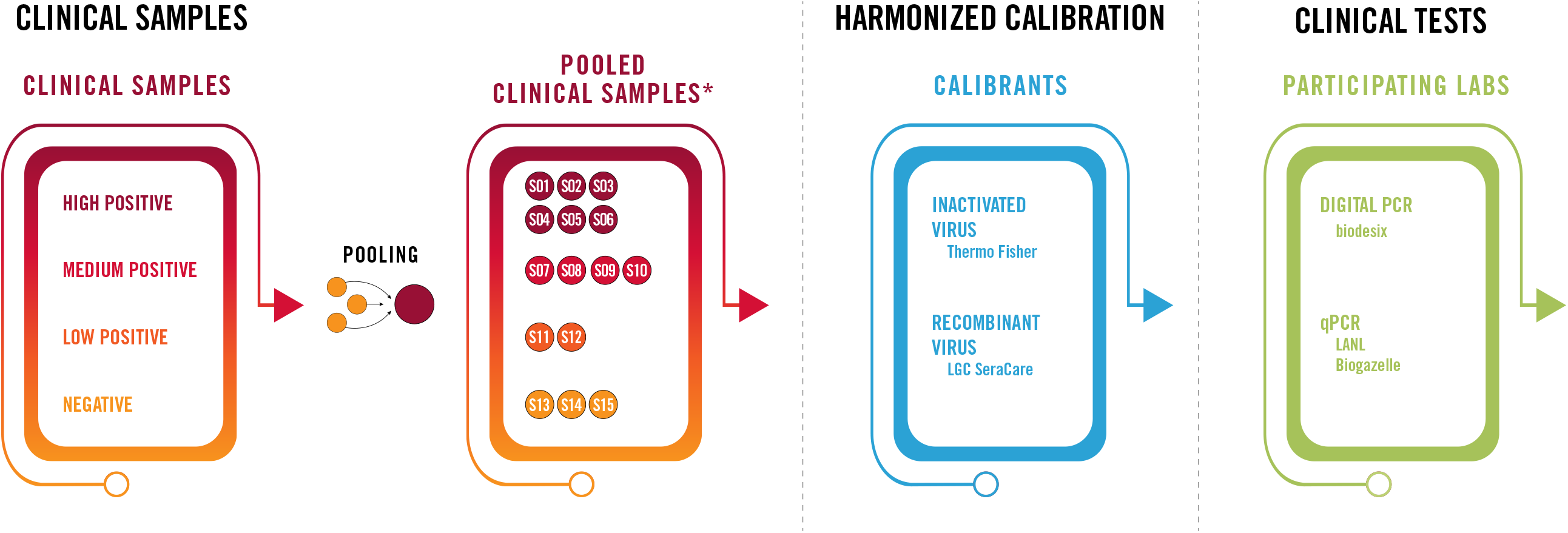
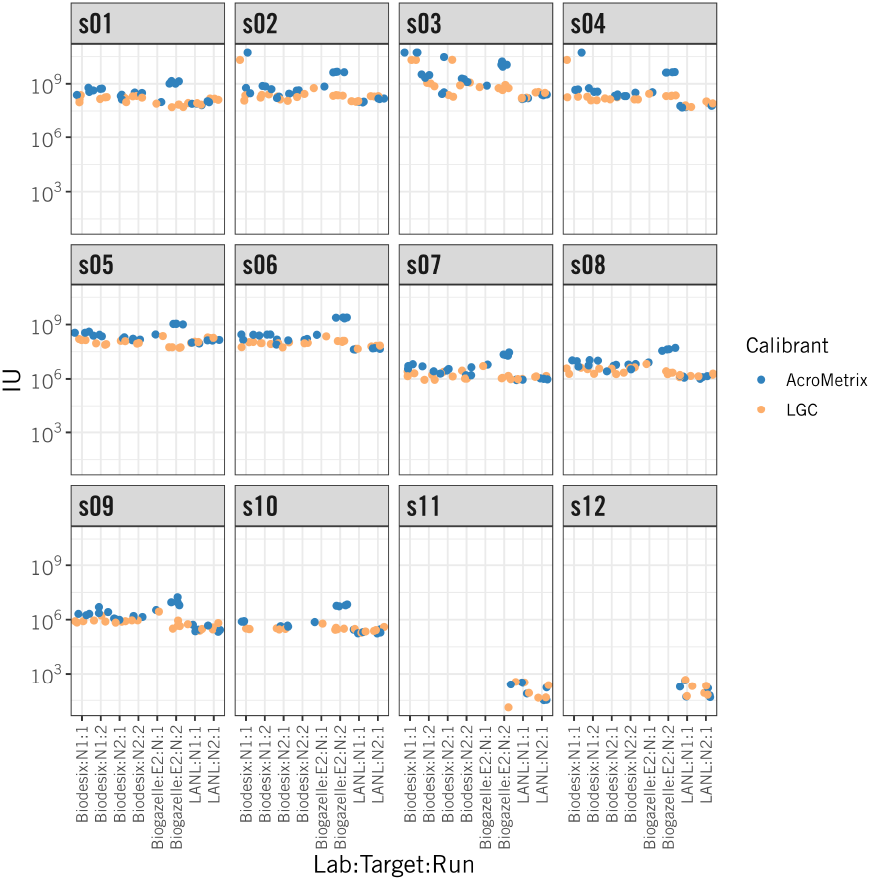
**(A)** Study design of a proof-of-concept application of the harmonized secondary reference materials to real clinical samples. 15 pools of biobanked samples (from 3 patients each) were aliquoted and shipped to three participating labs (Los Alamos National Laboratory (LANL), Biogazelle, and Biodesix), along with 3 calibrated standards representing the 3 classes: Inactivated virus: AcroMetrix/ThermoFisher Scientific, recombinant virus: AccuplexTM/LGC Clinical Diagnostics, and recombinant bacteriophage: Armored RNA Quant SARS-CoV- 2/Asuragen. Labs performed PCR using their described testing platforms and sent raw Cq or count data from their RT-qPCR or RT-dPCR platforms, respectively. **(B)** Harmonized measurements of viral load in 12 pooled clinical samples s01-s12: Calibrated measurements in clinical samples reported in IU mL^-1^. Pooled samples with different viral loads were measured by different labs along with two calibrant materials: LGC Clinical Diagnostics, and AcroMetrix. Data is shown after calibration of the raw data from both qPCR and dPCR platforms (Supplementary Figure 3).

### What we learned

It is practical and possible to harmonize standards and controls early in an outbreak or pandemic by strategically bringing together a diverse set of specialists. A thoughtful study design supported by clear SOPs and consistent data reporting make harmonization possible. Our CSWG experience cultivated a community of practitioners working collectively to respond to a global emergency, fostering new fruitful collaborations.

Our harmonization study and proof-of-concept study were conducted at different times, a year apart. We recommend that these studies be done simultaneously, with the same assays and lots of standards used in the harmonization. This improved design **(see “playbook” in Supplement 3)** can be periodically repeated to maintain comparability as the pathogens, assays and tests, sampling, and standards evolve.

The heterogeneity of laboratories, assays, and materials highlight the influence of these variables on test performance. The most significant artifacts we observed arose from standards developed from different materials. Full-genome inactivated virus standards and recombinant whole-virus samples behaved differently than the much smaller genomes with limited PCR target coverage of the bacteriophage materials. These different materials also had very different genome concentrations, requiring different dilution and handling in the lab.

The study design permits robust estimates of measurement uncertainty. Such estimates promise the ability to propagate uncertainties in calibrated results. This design ensures broad applicability to the heterogeneous tests and materials deployed internationally.

### How to do it next time

It is inevitable that new pathogens (viral, bacterial, fungal or parasites) will emerge. The WHO provides guidance on preparing secondary standards to test for emerging pathogens^10^. Our experience with the harmonization study conducted by the CSWG for SARS-CoV-2 is captured and presented **(Supplement 3)** in a generalized Playbook for Harmonization of Standards to Calibrate Pathogen Molecular Diagnostics.

### Summary

Both the harmonization and proof-of-concept studies demonstrate that it is possible to contribute to the standardization of measurement results for diagnostic laboratories through international and interdisciplinary cooperation. Our data provide a framework to break the glass when needed and rapidly harmonize molecular standards.

## Supporting information

Supplement 1: Recommended Standard Operating Procedure (SOP) for RT-qPCR for the CSWG Harmonization Study

Supplement 2: Recommended Standard Operating Procedure (SOP) for Proof-of-Concept Application of Harmonized Standards to Clinical Samples

Supplement 3: Playbook for harmonization of testing of emergent viral pathogens

## Data Availability

Raw data and scripts are publicly available in this Github Repository: https://github.com/msalit/harmonization_dashboard. Processed data can be visualized using the Shiny app: https://msalit.shinyapps.io/RNAstudy/.

https://msalit.shinyapps.io/RNAstudy/

## Acknowledgements

We acknowledge Lynne Sopchak for critically reading the manuscript and providing comments. We also acknowledge Andrew Hyatt for support with graphic design. Points of view in this document are those of the authors and do not necessarily represent the official position or policies of the U.S. Department of Commerce. Certain commercial software, instruments, and materials are identified in order to specify experimental procedures as completely as possible. In no case does such identification imply a recommendation or endorsement by the National Institute of Standards and Technology, nor does it imply that any of the materials, instruments, or equipment identified are necessarily the best available for the purpose. SS is supported by grants from the Chan Zuckerberg Biohub, San Francisco and the National Institute of Allergy and Infectious Diseases (NIAID).

## Figure Legends

**<u>Supplementary Figure 1:</u>** Calibration curves of each individual laboratory and assay using the 6-step dilution curves of the NIBSC 20/146 reference material. All data (RT-qPCR and RT-dPCR) have generated standard curves. Curve statistics are in Supplementary table 3.

**<u>Supplementary Figure 2:</u>** Expanded Box and whiskers represent median and interquartile ranges corresponding to the SARS-CoV-2 concentrations in IU, as depicted in Figure 1B in different types of standards: inactivated virus **(A)**, recombinant virus **(B)**, and bacteriophage **(C)**.

**<u>Supplementary Figure 3:</u>** Raw Cq values from LANL **(A)** and Biogazelle **(B)**, and raw counts from Biodesix **(C)** of the 15 pools of samples along with the secondary reference standards. Red points correspond to samples with undetectable amplification by PCR.

