## Supplement 1: Recommended Standard Operating Procedure (SOP) for RT-qPCR for the CSWG Harmonization Study for "A playbook for harmonization of standards for emergent viral pathogens"

CRITICAL NOTE: This SOP is not intended to be prescriptive, but to enable consistency of experiments across the participating laboratories. All process deviations must be recorded.

We recommend using existing lab procedures as implemented in your laboratory. Please plan to report this procedure (directly or by reference).

#### Objective:

Our Harmonization Study will establish the equivalence of SARS-CoV-2 RNA target concentrations across a panel of materials and calibrate those results against the candidate WHO International Standard (IS) reference sample. Those materials included in the study will have a basis to assert traceability of their viral RNA concentration levels to the value of the WHO IS. All data and results will be made publicly available without embargo as soon as results are validated. Following studies from the CSWG will employ these materials to develop benchmarking and validation kits.

This study is not a comparison of tests or labs. This study is not a survey of test or method performance for limit of detection, precision, or repeatability; it is not a study of commutability. This study will not evaluate material homogeneity or stability.

#### Description:

The study will analyze measurement results from approximately 10 established laboratories representing leading clinical labs, test developer labs, and national measurement institutes. These labs will measure a panel of approximately 10 different SARS-CoV-2 viral RNA preparations that are widely available and used as standards and controls. The preparations will include inactivated virus samples, recombinant virus samples, and recombinant bacteriophage samples, all suitable as full-process controls that can be extracted as a mock clinical sample. Measurements will include reverse transcription-quantitative real-time PCR (RT-qPCR) and reverse transcription-digital PCR (RT-dPCR) methods and will employ standards (e.g. MIQE) and best practices in reporting protocols, conditions, experimental factors, and results.

We will establish an open, public repository for the data, annotation, methods, and analysis of the study. The design of this resource will accommodate distribution of information to, as well as collection of the study questionnaires and results from the participants.

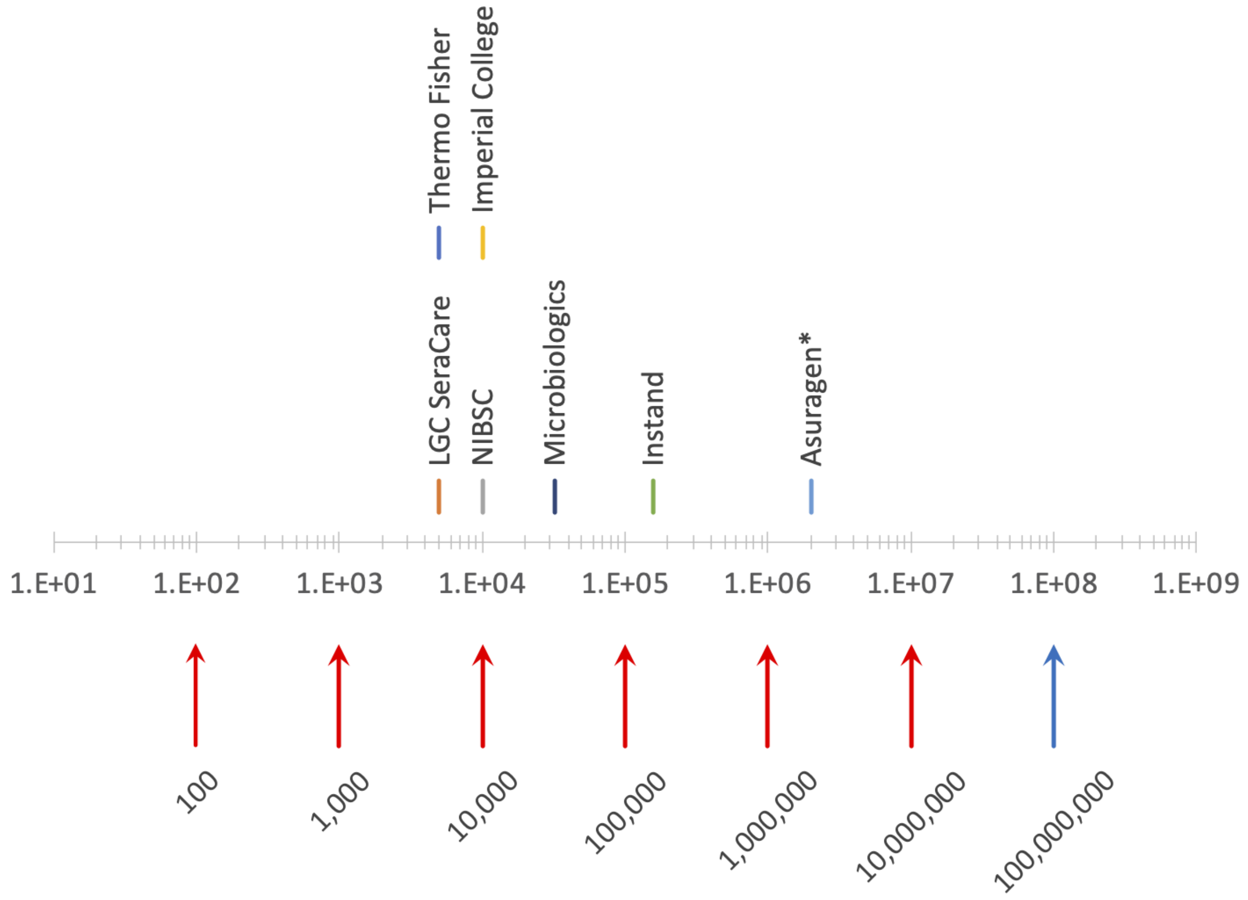


**Figure Caption:** This graphical representation of the nominal concentrations of the materials included in the Harmonization Study Panel is the guide we used to develop the recommended SOP below. All samples are at their nominal concentration except for the Asuragen sample, which is diluted 1:$10^4$ for use in the study. (copies/mL vs. IU)

### Experiment Design:

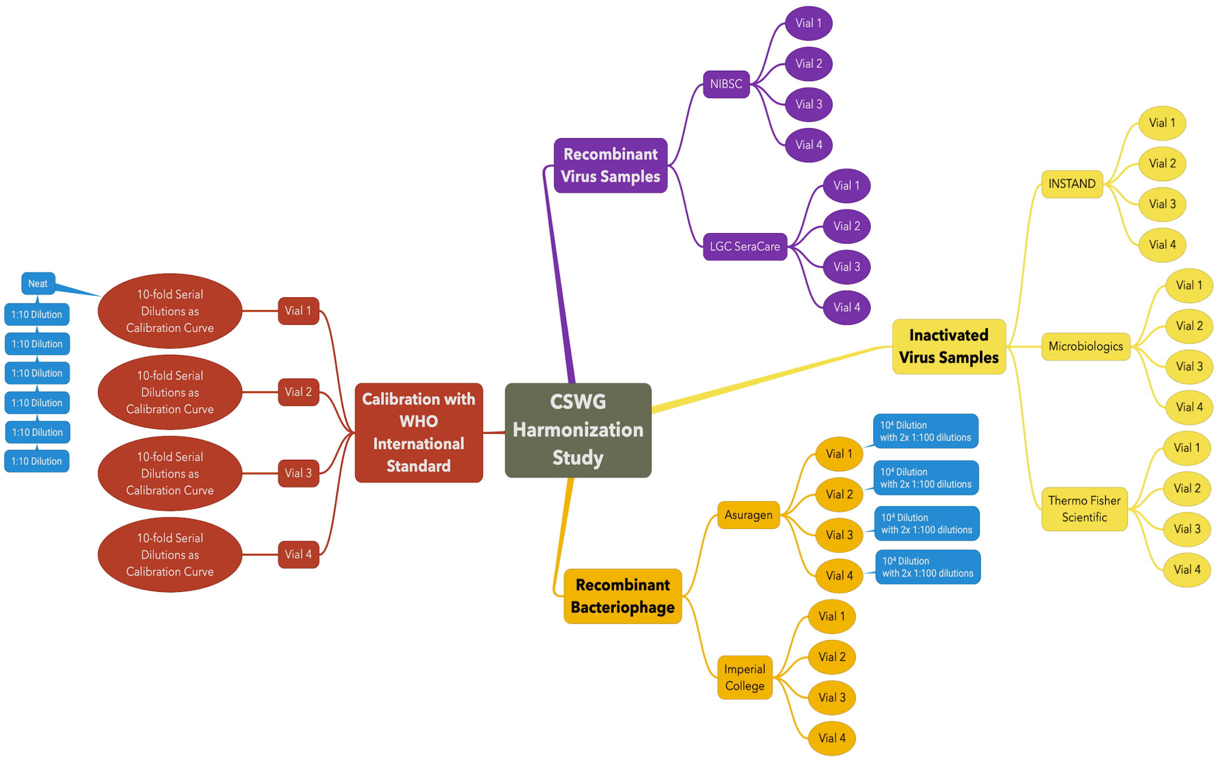


Participating laboratories will calibrate the 8 sample panel against the WHO International Standard (WHO-IS).

The diagram above describes the sample measurement strategy:

- There are 4 independent vials of the WHO-IS. Each vial of WHO-IS will be used to make an independent 7-point calibration curve by 6 serial 1:10 dilutions.
- Four vials of each of the 8 samples will be measured against this calibration curve. All vials will be measured as provided, EXCEPT for the Asuragen sample. Each vial of the Asuragen sample will be diluted 1:$10^4$ before measurement.

**Materials Provided:**

**WHO-IS:** 4 vials, each used to make an independent 7-point calibration curve, 1:10 serial dilution.

**7-panel standards (Inactivated Virus, Recombinant Virus, Recombinant Bacteriophage):** 4 vials of each to be measured against the WHO-IS calibration curve. All vials will be measured as provided, EXCEPT for the Asuragen sample, to be diluted 1:10,000 before measurement.

#### **Supplied Reagents:** Each lab will receive **4 vials** of each of the following:

- Instand; **Catalogue no. 340069.** Store at 4 °C.
- Microbiologics: Helix Elite Inactivated SARS-CoV-2 Whole Virus (Pellet); **Catalogue no. HE0065N.** Store at 4 °C. (more info)
- Thermofisher: AcroMetrix SARS-CoV-2 Control; **Catalogue no. 954517.** Store at -20 °C. (more info)
- LGC SeraCare: AccuPlexTM SARS-CoV-2; **Catalogue no. 0505-0168.** Store at 4 °C. (more info)
- NIBSC: SARS-CoV-2 RNA; **Catalogue no. 20/138.** Store at -20 °C . (more info)
- Asuragen: Armored RNA Quant SARS-CoV-2 Control; **Catalogue no. 52036.** Store at -20 °C. (more info)
- Imperial College: MS2-SARS-CoV-2 nucleocapsid gene Virus-like Particles; **non-commercial.** Store at -20°C. (more info)
- Zeptometrix: NATtrol™ SARS-CoV-2. **Catalogue no.** NATSARS(COV2)-ST. Store at 2-8 °C. (more info)
- WHO International Standard NIBSC; **NIBSC code: 20/146.** Store at -20°C. (more info)

#### General preparations:

1. Work in biosafety containment level 2 (BSL2).
2. For dry stocks (Instand and Microbiologics), centrifuge all standard tubes before opening, to capture any material stuck on the tube before resuspension.
3. Resuspend Instand stock in 1.1mL of bidi stilled water (for a final concentration of 157,000 copies/mL)
4. Resuspend Microbiologics (Helix Elite Inactivated SARS-CoV-2 Whole Virus) in 1mL in water or viral transport media (not supplied) for a final concentration of 3.2 X $10^4$ copies per mL

### Preparation of NIBSC WHO-IS dilution series:

5. Prepare the WHO-IS dilution series (NIBSC stock concentration is $10^8$ copies/mL). The purpose is to have 4 replicate standard curves. These dilutions should be prepared and used as soon as possible after preparation. Please take note of any storage times and conditions for these calibrants.
6. (Storage recommendation for the reconstituted WHO IS in case you need to go back)
   This is the recommended procedure to generate a standard curve. It will be done in quadruplicates from 4 independent vials of NIBSC stocks. At minimum, you should use template at directly from stock ($10^8$ copies/mL) for the upper limit, and the lowest should be $10^2$ copies/mL. Laboratories using dPCR approach should use the dilution series appropriate for their platform(s).

   We recommend a 10-fold dilution series before proceeding with RNA extraction or qPCR **(Figure 2)** covering the following 7-points to include (starting from native stock, then 10-fold step dilutions). **This dilution series applies to the NIBSC WHO-IS.** Note that this is an independent curve from each vial to generate 4 standard curves (each dilution step generated by adding 100µL of previous dilution + 900uL of water):

   $10^8$ copies/mL (directly from stock, no dilution)
   $10^7$ copies/mL: 1:10 from $10^8$ copies/mL
   $10^6$ copies/mL: 1:10 from $10^7$ copies/mL
   $10^5$ copies/mL: 1:10 from $10^6$ copies/mL
   $10^4$ copies/mL: 1:10 from $10^5$ copies/mL
   $10^3$ copies/mL: 1:10 from $10^4$ copies/mL
   $10^2$ copies/mL: 1:10 from $10^3$ copies/mL

7. You will have a total of 4 replicates X 7 concentrations = 28 tubes to generate the 4 standard curves. Record on the tube and *data entry form* which replicates are used for the dilutions (e.g. replicate 1, $10^8$ copies/mL, etc.):

### Preparation of other standards:

Goal: For each standard, you will record a single PCR measurement from each of the 4 independent vials. Indicate in the *Data Entry* form whether you will record Cq values from a quantitative PCR platform, or number of copies/mL from a digital PCR platform and record the metadata assay's characteristics in the *Google form.* This is a single form that each participating lab will submit once:

Participating lab:
Is there an RNA extraction step (Y/N):
RNA extraction platform:
PCR instrument:

PCR kit:
Digital or RT-qPCR:
Primer sequences if available: (or target gene if primer sequence is not available)
Data processing (e.g. procedure to generate number of copies):

8. For lyophilized standards (name the lyophilized standards), resuspend in nuclease free PCR grade water as indicated below:

9. Bring standards to room temperature.
10. All standards are intended to be used neat (i.e. directly from stock without dilution) except Asuragen (Armored RNA Quant SARS-CoV-2 Control).
    a. At the labs discretion any of the standard materials I1, I2, I3, R1, R2, P1,P2 can be diluted to increase the volume as needed for your assay. Dilute in media appropriate for the standard, as described in the table above or using your own Viral Transport Media (VTM). Please record and report any dilutions prior to measurement.
11. Dilute Asuragen stock ($2x10^{10}$ copies/mL) in two steps to generate a $2x10^{6}$ copies/mL dilution. **All dilutions need to be performed in TSMIII buffer, provided in the Asuragen Armored RNA Quant kit**:
    Step 1: dilute stock 1:100 as follows: 4uL of stock (Armored RNA Quant SARS-CoV-2 Control) + 396uL of TSMIII buffer. This is your **first intermediate dilution**.
    Step 2: dilute the 1:100 as follows: 4uL of first intermediate dilution at 1:100 + 396uL of TSMIII buffer. This is your **final** dilution for either the integrated qPCR workflow (combined extraction and qPCR), or separate RNA extraction and qPCR steps.
    Perform separate dilutions for each of the 4 stocks of Asuragen standard as described in steps 1-2 above.
    The concentration of this final diluted solution is $2x10^{6}$ copies/mL.

12. In addition to the 28 samples to establish standard curve, you will run each of these standards, **do not vortex to prevent shearing the material- only tap gently to mix,** you will run each in 4 replicates (8 standards X 4 replicates = 32 independent PCR reactions):

- Instand Concentration: 157,000 copies/mL
- Microbiologics Concentration: 32,000 copies/mL
- Thermo Fisher(AcroMetrix) Concentration: 5,000 copies/mL
- LGC SeraCare Concentration: 5,000 copies/mL
- NIBSC Concentration: 10,000 copies/mL
- Asuragen Concentration: $2x10^{6}$ copies/mL (1:10,000 from stock)
- Imperial College Concentration:10,000 copies/mL

13. Run 4 independent non-template control reactions (PCR-grade water).
14. In total, you will have 28 reactions (WHO-IS NIBSC standard dilutions) + 32 (standard replicates) + 4 non-template controls = 64 PCR reactions.
15. You are invited to add as many additional controls as you would like. The minimum we suggest including is the above 64 reactions.
16. Proceed with your lab's RT-qPCR or RT-dPCR protocol of your choice. You are only expected to run the assay on a single platform.

### Data Entry:

1. Note all the metadata into the data entry form (google form)
2. Template for including Cq or number of copies/mL (google sheet) per lab

#### General Notes:

- All data should be reported as raw and unprocessed as possible. The expectation is to do a single run per intended vial and report all data. If reactions fail or no amplification is detected, record that.
