## Supplement 2: Recommended Standard Operating Procedure (SOP) for Proof-of-Concept Application of Harmonized Standards to Clinical Samples for "A playbook for harmonization of standards for emergent viral pathogens"

CRITICAL NOTE: This SOP is not intended to be prescriptive, but to enable consistency of experiments across the participating laboratories. All process deviations must be recorded.

We recommend using existing lab procedures as implemented in your laboratory. Please plan to report this procedure (directly or by reference).

#### Objective:

This clinical study is an extension of the CSWG harmonization study to establish international units for common molecular standards for SARS-CoV-2 testing. The first part of the study established target concentrations across a panel of materials and calibrated those results against the candidate WHO International Standard (IS) reference sample. After calibrated international units (IU) have been assigned, this study will apply these standards in 3 clinical laboratories to clinical samples (frozen nasopharyngeal samples) spanning a range of different SARS-CoV-2 cycle threshold values.

Similarly to the first part of the harmonization study: This study is not a survey of test or method performance for limit of detection, precision, or repeatability; it is not a study of commutability. This study will not evaluate material homogeneity or stability. The primary goal of this section is to establish the reliability and reproducibility of inferring viral loads using calibrated samples run on different molecular testing platforms.

#### Description:

The study will measure the SARS-CoV-2 viral load using three molecular standards calibrated to the WHO NIBSC standards to estimate the quantity of the virus in the clinical samples using international units (IU). Ten separate pools of biobanked clinical samples from routine testing will be aliquoted, shipped and analyzed using 3 reverse transcription-quantitative real time PCR (RT-qPCR). The three standards will include an inactivated, recombinant virus and recombinant bacteriophage standards, which have already been assigned estimated IU/mL in the previous exercise. The 12 positive pooled samples will span a wide dynamic range to cover low, intermediate, and high viral loads based on the original quantification cycle ($C_q$) values obtained during initial testing and 3 negative pooled samples. Ideally, the ground truth on these quantitative values should be determined by a method distinct from methods used for calibration. Each laboratory will report their raw Cq (for RT-qPCR) or copy numbers (for RT-dPCR), as well as the extrapolated IU/mL values using the reference standards.

### Experimental Design:

Participating laboratories will measure 12 pools of SARS-CoV-2-positive and 3 SARS-CoV-2 negative samples, as well as 3 standards, each in duplicate measurements.

### Participating laboratories:

- LANL
- Biogazelle
- Biodesix

### Materials Provided (in tubes, 1mL each):

- Clinical samples (please confirm receipt of all; you will be blinded to which samples are positive and which are negative):

12 positive samples
3 negative samples

- Standards (subset of analytes included in the standards harmonization study)

1. ThermoFisher Scientific (Inactivated Virus): AcroMetrix SARS-CoV-2 Control; **Catalogue no. 954517.** Store at -20 °C. (more info). **Contact:** Thermofisher (ref: 954517, lot: 062101). Study coordinator will send 1 box of 5 tubes to each lab. You will need to run 3 replicates of a **single** tube.

2. LGC Clinical Diagnostics (Recombinant Virus): AccuPlexTM SARS-CoV-2; **Catalogue no. 0505-0168.** Store at 4 °C. (more info). **Contact:** LGC Clinical Diagnostics; (ref: 0505-0159, lot: 1057651). Study coordinator will send 1 box of 5 positive and 5 negative vials. You will need to run 3 replicates of a **single** positive tube. If you can, add 3 replicates of a **single** negative tube, but this is optional.

3. Asuragen (Recombinant Bacteriophage): Armored RNA Quant SARS-CoV-2 Control; **Catalogue no. 52036.** Store at -20 °C. (more info). **Contact:** Asuragen; (ref: 52036C, DOM: 2021-10-01). Study coordinator will send 2 tubes of the standard. You will need to run 3 replicates of a **single** tube. The second tube is a backup; you only need to run 3 replicates from a single tube.

Note: you will be blinded to which of the 15 samples are positive or negative.

### General Preparations:

1. Receiving samples:  Document sample quality and confirm all samples are received by email (confirm receipt to study coordinator).
2. Work in biosafety level 2 (BSL2).
3. Follow your routine lab process for analyzing clinical samples: extraction, RT-qPCR. Document the qPCR platform, process, and report it along with the results.
4. Data entry: enter raw Cq values for each sample here:

| Sample number | Replicate 1 | Replicate 2 | Replicate 3 |
|---|---|---|---|
| 1 | | | |
| 2 | | | |
| 3 | | | |
| 4 | | | |
| 5 | | | |
| 6 | | | |
| 7 | | | |
| 8 | | | |
| 9 | | | |
| 10 | | | |
| 11 | | | |
| 12 | | | |
| 13 | | | |
| 14 | | | |
| 15 | | | |
| Thermofisher | | | |
| LGC Clinical Diagnostics | | | |
| Asuragen | | | |
