## Supplement 3: Playbook for harmonization of testing of emergent viral pathogens for "A playbook for harmonization of standards for emergent viral pathogens"

Public health response to pandemics needs diagnostic test results that can be compared across space and time. Regardless of whether the diagnostic data are qualitative (as used to determine "clinical positive/clinical negative") or quantitative (to establish a comparable measure of "viral load"), those data must be reported on a common scale.

We offer this "playbook" as a generally applicable tool to develop and conduct a harmonization study that establishes a reference scale of calibration standards and controls for diagnostic molecular testing of emerging pathogens. The playbook is based on the experience of the Coronavirus Standards Working Group (CSWG), who developed and conducted a harmonization study for SARS-CoV-2 molecular testing. We describe here the principles and key design factors for designing the study, selecting materials, selecting laboratories, validating the utility of calibration with pooled clinical samples, and analyzing and reporting the data (**see Playbook Figure 1**). Recommendations for conducting the study are included to make it a "plug-and-play" process. Our experience recommends that a Working Group (WG) be formed to conduct such a study[1]. This playbook is a recipe to break the glass when needed to rapidly harmonize molecular standards.

Standards harmonized using this approach will be reliable, platform-agnostic, and demonstrated to be clinically useful. Clinical results from calibrated tests will have a "y-axis" of pathogen units that can be compared to each other, enabling interpretation of pathogen load and transmission dynamics across studies. When International Standards (IS), such as those from the World Health Organization (WHO) are available, the values of calibration materials can be made traceable to the value of the IS[2,3].

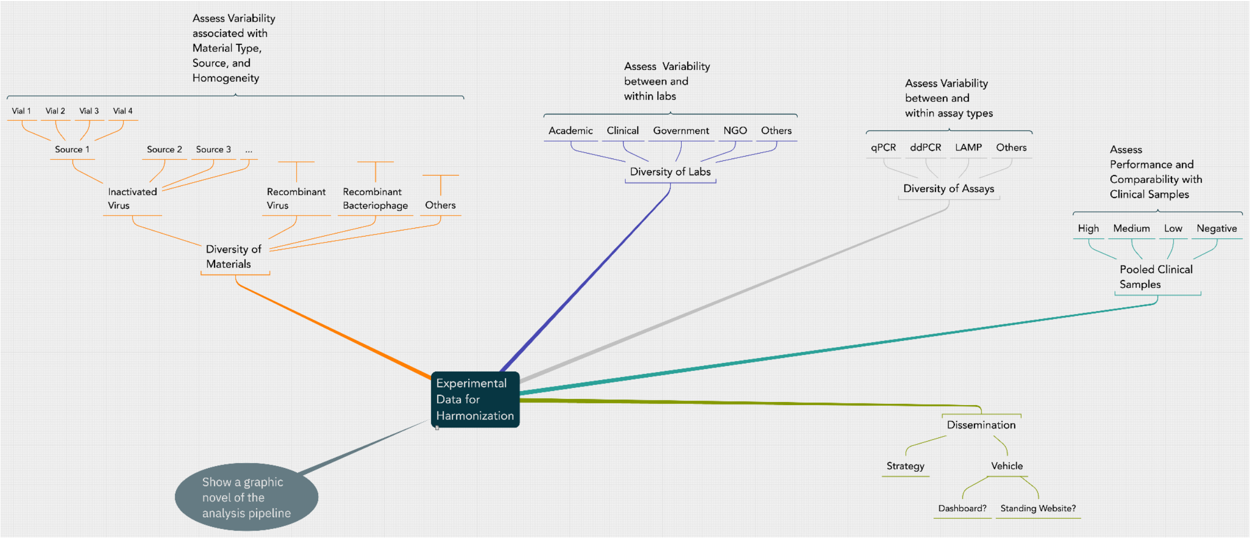

***Playbook Figure 1.*** *This figure shows the elements of the harmonization study recommended in this 'Playbook' and highlights the diversity of the materials, laboratories, and test methods that will contribute to the dispersion of measurement results.*

### Principles and Key Design Factors

#### Harmonization Study Design

Harmonization is based on an interlaboratory study that compares a set of representative calibration materials across multiple laboratories representing a sample of the population of testing laboratories. The recommended experiment design has replication sufficient to identify and assess sources of variability associated with sampling, handling, and technology platforms. This design is depicted in **Playbook Figure 1.** The design must be realized and described in a standard operating procedure (SOP) to assure that participating laboratories conduct experiments and report data consistently.

The harmonization study recommended in this playbook is not to compare tests or laboratories. The harmonization study is not a survey of test or method performance for limit of detection, precision, or repeatability; it is not a study of commutability. This study will not evaluate material homogeneity or stability.

All scientists and organizations participating in the WG shall consent in advance of the study composition to transparent, open, and unembargoed data sharing.

#### Establishing a Coordinating Laboratory

A successful harmonization study calls for a coordinating laboratory to act as a single point of logistical and operational leadership. The coordinating laboratory should be appointed by the WG to conduct study development and operation, and should have the remit to:

- convene the working group to establish the study
- lead the collective effort to select materials and measurement laboratories
- lead the work to develop and distribute the SOP
- develop the library of standards to be harmonized
- coordinate development of the pooled clinical samples
- assemble and distribute "kits" of all the materials to the measurement laboratories in a consistent manner
- establish a repository to collect the data
- coordinate data analysis

Where possible, the coordinating laboratory should have a documented and accredited quality system and use it to assure quality of the development of the SOP, the pooled clinical samples, development of the kits, and their distribution. Where a separate laboratory is used to develop the pooled clinical samples, the coordinating laboratory should assure appropriate documentation and oversight.

#### Selecting Calibration Materials to Harmonize

Representative materials that act as good surrogates for the pandemic pathogen should be selected from those that are widely available. For example, the set of materials might include different forms, such as inactivated virus, recombinant virus, recombinant phage, and possibly other relevant formulations. The most useful materials to include will be those that:

- can be used as mimics in the workflow as applied to clinical samples
- "widely available" calibration materials should be those distributed globally
- prefer materials that can be used without restriction
- prefer materials that have a path to persistent/enduring/ongoing availability
- prefer materials from a variety of sources, including academic and commercial laboratories

#### Selecting Laboratories to Provide Results for the Harmonization Measurements

The WG and the coordinating laboratory should recruit a population of laboratories to participate in the study. These laboratories should have well-established assays in place for the pathogen in clinical samples or be laboratories with special attributes that have well-established capabilities to contribute to the harmonization study (see list below for suggestions). All laboratories should have appropriate quality measures in place to control their assays, including experience with quality systems and analytical validation such as would be needed for accreditation.

The study laboratories should be drawn from a population of:

- Clinical testing laboratories (including ad hoc clinical laboratories)
- Reference laboratories (including National Metrology Institutes)
- Non-governmental organizations (such as WHO)
- Government and regulatory bodies overseeing pandemic response testing
- Professional society-designated laboratories
- Laboratories or organizations conducting proficiency tests
- Testing technology developers
- Laboratories from the affected areas, and underrepresented communities

#### Demonstrating that Harmonized Calibration Materials Harmonize Results with Clinical Samples

An experiment should be conducted in parallel with the harmonization measurements to demonstrate the relevance of the harmonization study and the utility of the harmonized calibration materials. The playbook suggests measuring a set of pooled clinical samples along with the calibration materials. Post-hoc analysis will provide calibrate results for the pooled clinical samples, and enable evaluation of cross-platform, cross-calibration material harmony. We recommend pooling clinical samples in tranches across the dynamic range of clinically relevant viral load to make a set of surrogates that can be distributed amongst the participating laboratories along with the calibration materials. Pooling clinical samples for this "proof of concept" element of the study will assure adequate volumes for distribution across participating laboratories.

Demonstrating that results from clinical surrogates are equivalent regardless of laboratory, platform, or calibration materials is the strongest way to show successful harmonization and calibration.

#### Data Analysis and Dissemination of Results

All data and analysis from the study should be open, public, unembargoed, and comply with FAIR [Findable, Accessible, Interoperable, and Reusable] Data Principles[7]. Transparency and analysis are facilitated by using a common template for raw data submission, and an open-source, publicly accessible analysis platform that uses a documented analysis strategy.

These open data and analysis tools will permit re-analysis and reuse of the data. Transparency and well-documented processes build confidence in the WG effort and the conduct of the study, leading to confidence in the results.

##### Analysis Description

We recommend using robust estimates for the calibrant values and their dispersions, using a Median as the value and the Median Absolute Deviation (MAD) as a measure of the dispersion. We recommend presenting the dispersion as twice (2x) the MAD as an approximation to a 95% confidence interval. Depending on the distribution of the results, other studies may use other appropriate statistical analysis methods.

- robust estimates of median and dispersion
- comparison of study estimates to nominal values of standards
    - copy numbers from providers, corrected to IS value

#### Harmonization Study Deliverables

The harmonization study should be published in the open literature, using a preprint server as appropriate to disseminate the study and results in a timely manner. Data availability is described above, and the study should be widely described in talks and on social media and other vehicles as it is conducted. The analysis tool should be made widely accessible as a public, web-hosted 'dashboard.' This can serve as a central repository of data, analysis, and results (see GitHub from CSWG Harmonization study). This repository will serve as a FAIR data repository.

We recommend the following as a statement of metrological traceability to be suggested to be included in the Certificate of Analysis accompanying calibration materials included in this study:

- "The quantitative value assigned to this material is traceable to the International Unit through the harmonization study conducted by the XX Pathogen Working Group on DD/MM/YYYY"

##### Bridging studies

The WG should encourage bridging studies for new calibration materials or new lots of existing materials. Such studies should include replication to assess dispersion, and we recommend validating the bridging study with inclusion of other harmonized calibration materials to verify their consistency as a quality check.

#### Generic Standard Operating Procedure (SOP) of Harmonization Study and Application on Clinical Samples to Detect Viral Nucleic Acid

A standard Operating Procedure (SOP) shall be distributed by the coordinating laboratory to all participating laboratories. The SOP is not intended to be prescriptive, but to enable consistency of experiments across the participating laboratories. All process deviations must be recorded. The laboratories shall use their existing laboratory procedures as implemented for testing (i.e. sample handling procedure). The participating laboratories should report this procedure (directly or by reference). All measurement results should be reported in the data reporting template developed and distributed by the Coordinating Laboratory. All measurements shall be done with at least quadruplicate replicates to robustly capture dispersion of measurements in each test.

##### Objective:

This Harmonization Study will establish the equivalence of measuring abundance of nucleic acid from novel virus targets in a range of concentrations represented from a panel of different materials from different sources, calibrating those abundances to a scale established by a selected reference, preferably an International Standard (IS) reference sample. Materials included in the study will have a basis to assert traceability of their viral nucleic acid concentration levels to the value of the IS. All data and results will be made publicly available without embargo as soon as results are validated. Following studies from the working group can employ these materials to develop benchmarking and validation kits.

This study is not a comparison of tests or laboratories. This study is not a survey of test or method performance for limit of detection, precision, or repeatability; it is not a study of commutability. This study will not evaluate material homogeneity or stability.

##### Description:

The study will analyze measurement results from approximately 5-10 established laboratories representing leading clinical laboratories, test developer laboratories, and national measurement institutes. These laboratories will measure a panel of approximately 5-10 different preparations of viral nucleic acid materials that are widely available and used as standards and controls. The preparations will include inactivated organisms, pathogen nucleic acid or synthetic viral genomes (DNA or RNA), which would be suitable as full-process controls that can be extracted as a mock clinical sample. Measurements will include nucleic acid amplification methods such as quantitative real-time PCR (RT-qPCR or reverse transcription-qPCR (RT-qPCR) and digital PCR (dPCR), and reverse transcription-dPCR (RT-dPCR) methods, and will employ standards, e.g. Minimum Information for the Publication of Quantitative Real-Time PCR Experiments (MIQE)[6], and best practices in reporting protocols, conditions, experimental factors, and results.

This harmonization study should also apply these standards to clinical samples of different viral loads: negative, low, medium, and high.

An open, public repository for the data, annotation, methods, and analysis of the study will be established with the involvement of expert ethics and legal consultant. The design of this resource will accommodate distribution of information to, as well as collection of the study questionnaires and results from the participants.

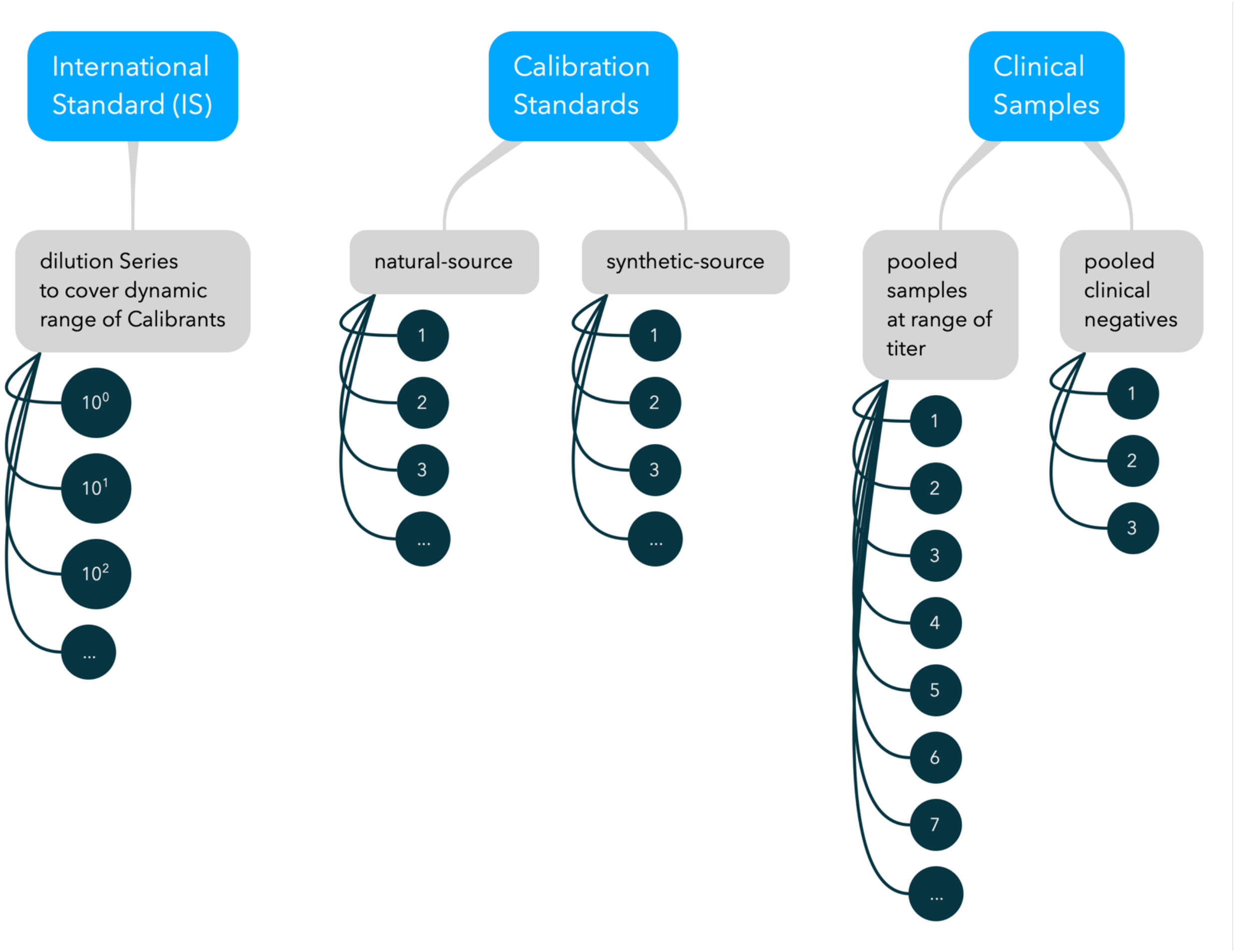


***Playbook Figure 2:*** *Samples in the harmonization study. Study design includes replication of each sample at each laboratory, not shown (4 vials are recommended of each standard and calibration material). Measurement assay replicates are not shown (we recommend 4 replicates per sample to robustly assess dispersion of each measurement).*

#### Experiment Design:

Participating laboratories will calibrate the panel of standards against the WG agreed upon International Standard (IS) for the measurement of the pathogen of interest. The diagram in **Figure 2** above describes the samples in the study.

The participating laboratories should follow the procedure outlined below:

- Each laboratory should follow the existing procedures for their testing assays, including positive and negative controls and quality measures to ensure assay reliability.

- The SOP should clearly list all material sent from the coordinating laboratory to the participating laboratories in a tabulated format. This should include: the name of the standard, the catalog number, storage information, diluent buffers for any necessary dilutions. Attach the manufacturer's instructions for use and any supporting documentation or citations.
- The SOP should state any necessary reagents for resuspending or diluting the IS, the standard material, and the clinical samples.
- Provide 4 independent vials of each sample, including the IS. Each vial of IS will be used to make an independent calibration curve by a dilution series. The dynamic range covered by the dilution series should accommodate the concentrations of all the standard materials selected for the harmonization study (5–10-point dilution series).
- Clinical samples should include replicates from pooled samples covering a range of values: negative, low, medium, and high viral loads based on an independent assay not included in the harmonization study (e.g. viral titers based on plaque assays). Identical vials from the pooled clinical samples should be shipped from the coordinating to the participating laboratories.
- For all samples, each participating laboratory should confirm receipt of all samples in good condition, and communicate any issues immediately to the coordinating laboratories, e.g. missing vials or samples thawing during shipment.
- Clearly state the biosafety containment level and any personal protective equipment necessary to handle the samples and conduct the measurements.
- Laboratories should follow their own assay-specific SOPs for qPCR, dPCR, or other nucleic acid measurement assays.
- Each of the participating laboratories should fill a standardized *data entry form* to record the raw measurements from each replicate of the IS standard dilution series, the standard materials, the clinical samples and any controls, including non-templated controls. If a participating laboratory uses multiple assays for measurement, they should fill a separate data entry form for each assay used in the harmonization study.
- Each of the participating laboratories should record the metadata assay's characteristics in the *metadata form.* If a participating laboratory uses multiple assays for measurement, they should fill a separate metadata form for each assay used in the harmonization study.
- The participating laboratories should send a dated copy of the data entry forms and metadata forms to the coordinating laboratories, along with clear notes on any issues arising during the experiment.
- The WG should convene regularly to ensure timely delivery, review and analysis of the data to the analysis laboratory determined by the coordinating laboratory in consultation with the WG.
