## Supplement 4: Online Methods: for "A playbook for harmonization of standards for emergent viral pathogens"

**Study overview:**

The goal of this harmonization study was to establish the equivalence of SARS-CoV-2 RNA target concentrations across a panel of materials and calibrate those results against the candidate WHO International Standard (IS) reference sample. Those materials included in the study will have a basis to assert traceability of their viral RNA concentration levels to the value of the WHO IS. All data and results are publicly available without embargo as soon as results are validated (Shiny app: https://msalit.shinyapps.io/RNAstudy/). Following studies from the CSWG will employ these materials to develop benchmarking and validation kits. This study is not a comparison of tests or laboratories. This study is not a survey of test or method performance for limit of detection, precision, or repeatability; it is not a study of commutability. This study will not evaluate material homogeneity or stability. Participating laboratories calibrated an 8 secondary reference material sample panel against the WHO IS: NIBSC 20/146. There were 4 independent vials of the WHO-IS. The labs used each vial of WHO-IS to make an independent 7-point calibration curve by 6 serial 1:10 dilutions. Four vials of each of the 8 samples were measured against this calibration curve. All vials were measured as provided, except for the Asuragen sample. Each vial of the Asuragen sample was diluted 1:$10^4$ before measurement. Reference materials from Asuragen were aliquoted at the coordinating lab (Stanford) and distributed to all participating laboratories, then diluted in TSMIII storage buffer (Asuragen Inc.) to a final 1:10,000 dilution from stock concentration. The laboratories received two standard operating procedures: one for reverse transcription-quantitative real-time (RT-qPCR) and the second for reverse transcription digital PCR (RT-dPCR) describing the materials, dilution series, as well as data and metadata upload links.

### Acquisition of secondary reference materials

The reference material included 4 inactivated virus samples (Instand, Microbiologics, Thermo, and Zeptometrix), 2 recombinant virus samples (LGC Clinical Diagnostics, and NIBSC), and recombinant bacteriophage samples (Asuragen, and Imperial College), all suitable as full-process controls that can be extracted as a mock clinical sample. The material descriptions are listed in detail in **Supplementary Table 1A.** Measurements included RT-qPCR and RT-dPCR methods and employed standards (e.g. Minimum Information for Publication of Quantitative (MIQE) Real-Time PCR experiments or Digital MIQE Guidelines) and best practices in reporting protocols, conditions, experimental factors, and results. Each laboratory employed their existing protocols, RNA extraction or no extraction and a 1-step or 2-step RT-qPCR or RT-dPCR. The laboratories and PCR protocols used are listed in **Supplementary Table 1B.**

### Participating laboratories and local testing protocols

1. Biodesix:

This lab employed the Bio-Rad QX200 digital PCR (dPCR) instrument, using the Bio-Rad SARS-CoV-2 ddPCR Kit (Part No 12013743). The lab used 100µL of sample for RNA extraction using the Quick-RNA Viral 96 Kit (Zymo Research), then 13.8 % of the RNA eluate was input into the RT-dPCR reaction. Target values are reported as total copies detected per dPCR reaction. The lab tested 2 SARS-CoV-2 targets: N1, N2 and a reference RPP30 target, primer sequences in CAPS:

N1:

GACCCCAAAATCAGCGAAATgcaccccgcattacgtttggtggaccctCAGATTCAACTGGCAGTAACCAGA.

N2: TTACAAACATTGGCCGCAAAttgcacaatttgcccccagcgcttcagcgTTCTTCGGAATGTCGCGC.

RPP30:

AGATTTGGACCTGCGAGCGggttctgacctgaaggctctgcgcggactGAGCGGCTGTCTCCACAAGT.

All samples were processed in a single batch, except for Zeptometrix (received at a later date).

2. Biogazelle:

The lab extracted RNA using the PurePrep 96 Nucleic Acid Purification System (magnetic bead extraction); cat.nr: OE-17060-00. Kit used: PurePrep Pathogen kit (10*96); catalog number: OE00290960. RT-qPCR reactions were performed on the Bio-Rad CFX 384 Touch Real-Time PCR Detection System and analyzed using the One Step PrimeScript III RT-PCR Kit (Takara); catalog number: RR600.

Two targets were used to detect SARS-CoV-2 in the FAM channel:

Target 1=E-gene:

E_Sarbeco_F ACAGGTACGTTAATAGTTAATAGCGT

E_Sarbeco_P1 FAM/ZEN-ACACTAGCCATCCTTACTGCGCTTCG-IBFQ

E_Sarbeco_R ATATTGCAGCAGTACGCACACA

Target 2=N2-gene:

N2-CDC-F TTA CAA ACA TTG GCC GCA AA

N2-CDC-P FAM/ZEN-ACA ATT TGC CCC CAG CGC TTC AG-IBFQ

N2-CDC-R CG CGA CAT TCC GAA GAA

The lab also used an internal spike-in control (SIC) to test successful amplification, which is measured in the HEX channel:

SIC_F TCAGAGCCTAATCAGTTCAG

SIC_P GCCCTCGGCCTACATGT

SIC_R GAGTATGCGTATTACCCGTA

3. Bio-Rad:

All samples were extracted together on a Kingfisher with the Thermo Fisher Scientific MagMAX Viral/Pathogen Nucleic Acid Isolation Kit. All samples were then batch analyzed on a single dPCR plate (Batch 1). A second follow-up dPCR run (Batch 2) containing (1) WHO-IS Vial 4 Level 2, (2) all four Asuragen sample eluates further diluted by a factor of 10, (3) all four NIBSC sample eluates diluted by factors of 10 and 100, and (4) all four Imperial College unextracted samples. The dPCR reactions were run on the Bio-Rad QX200 AutoDG Droplet Digital PCR System (Part No 1864100), Bio-Rad C1000 Thermocycler (Part No 1851197), using the Bio-Rad SARS-CoV-2 ddPCR Kit (Part No 12013743), all according to the manufacturers' instructions. The primers and probes sequences are the same as those used in the CDC 2019-nCoV Real-Time RT-PCR Kit, as described above for Biodesix. dPCR data produces distinct clusters for each combination of positive target signal (e.g. N1+ N2- RP-, N1+ N2+ RP-, and N1+ N2+ RP+ are three distinct clusters). Clusters are manually identified by the user based on their relative positions and their appearance in the positive control (SARS-CoV-2 RNA transcripts and Human RNase P human genomic DNA). Following thresholding, the analysis software directly quantitates the copy concentration (cp/uL) of each dPCR reaction well by applying a Poisson statistical model. Droplet digital PCR is directly quantitative by nature and no standard curve is required.

4. LabCorp:

RNA extraction was performed using the Thermo Fisher Scientific MagMAX™ Viral/Pathogen Nucleic Acid Isolation Kit, A48383. RT-qPCR was performed on the QuantStudio™ 7 Flex using the Applied Biosystems™ TaqPath™ 1-Step Multiplex Master Mix, NO ROX: A28523; with a laboratory developed test primer mix, which received emergency use authorization.

5. Los Alamos National Laboratory (LANL):

LANL performed a 2-step RT-qPCR, with RNA extraction performed using the Thermo Fisher MagMAX Viral/Pathogen Nucleic Acid Isolation Kit, A48383. RT-qPCR was performed and data processing conducted on the Applied Biosystems™ 7500 Fast Dx Real-Time PCR Instrument using a modification of the US CDC assay.

6. Massachusetts Consortium on Pathogen Readiness (MassCPR):

The Massachusetts General Hospital performed the test on the Fluxergy's COVID-19-Sample-to-Answer-RT-PCR test, designed for processing, amplifying, and detecting SARS-CoV-2 from nasopharyngeal (NP) swab samples in Viral Transport Media. The Fluxergy analyzer is an RT-qPCR device that uses a test card with 6 wells, each with a unique Cq value, to determine the presence of SARS-CoV-2 virus in a single reaction. The test is designed to provide a qualitative positive result if at least two wells have Cq values $< 45$ cycles, otherwise the result is negative. The amplification uses FAM probes attached to forward and reverse primers to amplify SARS-CoV-2 N-gene and polyprotein gene, ORF1ab. Extraction process takes place in the test card during direct qPCR reaction, thus manual extraction of the RNA is not required. An internal (spiked) control, MS2ic3, amplified using a Texas Red Probe, is used in the background of every test to validate a true negative. The Fluxergy system is semi-quantitative and consists of Fluxergy analyzer, Fluxergy test card and Fluxergy reaction mixture and Fluxergy Works software. We performed the analysis according to the manufacturer's instruction manual. We prepared the samples by either diluting or thawing as required in accordance with the SOP. We thawed samples and pipetted 14 µL of sample to the reaction mixture tube (126 µL) and mixed well with the pipette. We scanned the barcode on individual test card package and typed the test name and sample specific information in the notes section. If a test was a retest, we named the test '_Retest'. Next, we removed the test card from the package and placed it on a flat surface. We immediately pipetted 130 µL of master-mix (sample and reaction mixture) into the loading port of the test card. After loading, we capped the loading port loaded the card in the correct orientation. We ran the test within 5 minutes of mixing the reaction, then ran the tests, which took 1 hour to complete, then exported the results into an excel file. Finally, we took an average of Cq value from all wells that had a Cq value < 45 to get a single Cq value for each sample.

7. Mayo Clinic:

The lab processed all the samples for RT-qPCR in a single batch. The samples were analyzed on the cobas 6800 and cobas SARS-CoV-2 assay (Roche Molecular Systems, Inc, Branchburg, NJ). The test is intended for the qualitative detection of nucleic acids from SARS-CoV-2. Using target-specific primers and probes, this assay amplifies and detects both the ORF1a (nonstructural protein) sequence of SARS-CoV-2 and the E gene (envelope protein) sequence common to the subgenus Sarbecovirus.

8. Medical University of South Carolina (MUSC):

The lab used a 2-step RT-qPCR analysis system. The samples were processed in one batch using the Abbott Alinity m System and Abbott Alinity m SARS-CoV-2 AMP kit. The PCR protocol detects a single cycle threshold value targeting RNA-dependent RNA polymerase and nucleocapsid (RdRP:N) genes.

9. National Institute of Biology (NIB):

NIB used a 1step RT digital PCR (RT-dPCR) analysis system. RNA was extracted using the QIAamp Viral RNA Mini Kit (Qiagen) according to the manufacturer’s instructions. PCR reactions were performed in T100 thermal cycler (Bio-Rad) using CDC N2 assay and the One Step RT-ddPCR Kit (Bio-Rad) according to the manufacturers’ instructions. Partitions were read out and analyzed using QX200 Droplet Reader (Bio-Rad) and QuantaSoft software (Bio-Rad).

Samples were analyzed in 3 batches for each group of 9 samples, where the first batch included a single repeat of all 9 samples, followed by batches 2 and 3, and so forth. Thresholds to determine positive samples were previously established using negative and positive samples.

10. National Institute for Standards and Technology (NIST):

NIST performed RT-dPCR, and hence no dilutions of the WHO IS were performed. The lab performed RNA extractions on each type of standard/material each day, on 3 different days. The extracted RNA material was split into 3 technical replicates for RT-dPCR each day, resulting in 9 total replicates. RNA was extracted using the QIAamp Viral RNA Mini Kit (QIAGEN, catalog number: 52906). Digital PCR reactions were performed on the Bio-Rad QX200 AutoDG Droplet Digital PCR System using the One Step RT-ddPCR Kit (Bio-Rad) according to the manufacturers' instructions. For ease of processing, the lab diluted the WHO IS and the NIBSC working reagent 1:100 before extraction (in addition to diluting the Asuragen reagent). The pre-PCR dilution factors, as well as extraction concentration/dilution factors, were all adjusted for the reported values.

Primers and probes: U.S. CDC N2 Probe= FAM-ACA ATT TGC CCC CAG CGC TTC AG-(MGB) F=TTA CAA ACA TTG GCC GCA AA; R=GCG CGA CAT TCC GAA GAA

11. UK National Measurement Laboratory (NML) LGC:

The NML lab performed RNA extractions using QIAamp Viral RNA Mini Kit (Qiagen). Digital PCR reactions were then performed using the Bio-Rad QX200 AutoDG Droplet Digital PCR System, and One-Step RT-ddPCR Advanced Kit for Probes (Bio-R ad), with the CDC N2 assay (position GenBank MN908947.3 position 29164-29230). Quality checks were performed of the dPCR data. Data compared to a positive dPCR control to ensure dPCR was performing. Thresholding was performed automatically across all wells in an experiment by the instrument and checked if appropriate (software version QuantaSoft 1.7.4.0917). Calculate the lambda value from the number of positive and accepted droplet numbers. Converted this into final concentration copies/uL in a reaction, using partition volume for the dPCR and reaction volume. This was then converted in copies/µL of elute factoring in the template volume added to the reaction. Factoring in the volume extracted we calculated copies/mL of the original unextracted sample. These raw values were uploaded to the CSWG central data repository.

12. Quest Diagnostics:

Molecular analysis at the Quest laboratory utilized the Quest Diagnostics SARS-CoV-2 RNA Qualitative Real-Time RT-PCR emergency use authorization (EUA) assay, performed according to the package instructions for use (FDA: https://www.fda.gov/media/136231/download; accessed on 11 April 2024). A total of 200 µL of patient specimen is combined with 250 µL of lysis buffer in the first step of the nucleic acid isolation procedure. Briefly, the assay utilizes a one-step reverse transcription and PCR amplification with SARS-CoV-2 specific primers and real-time detection with SARS-CoV-2 specific probes for the N1 and N3 targets of the virus nucleocapsid gene. Following RT-PCR, a specimen is deemed positive for SARS-CoV-2 RNA when the Cq values for both targets are less than 40 cycles. A specimen is deemed negative when the Cq values for both targets are ≥40 cycles and the internal amplification control is valid. If the Cq value for a single detector is <40 cycles while the Cq for the second detector is ≥40 cycles, the test result is deemed inconclusive, and the specimen is then retested.

13. Stanford Medicine:

The Stanford laboratory analyzed all samples in a single batch. RNA extraction was performed using the Janus G3 Reformatter and Chemagic™ 360 using Chemagic™ Viral DNA/RNA 300 Kit (PerkinElmer®, Catalog No. CMG-1033-S). RT-qPCR was performed using the Applied Biosystems™ QuantStudio™ 7 Pro Real-Time PCR System, with the PerkinElmer® New

Coronavirus Nucleic Acid Detection Kit (2019-nCoV-PCR-AUS). The primers target the Nucleocapsid (N) and Orf1ab genes. Laboratory steps have been performed according to the developer's instructions included in the FDA Emergency Use Authorization document: https://www.fda.gov/media/136410/download.

14. Western University (WesternU) of Health Sciences:

Western University of Health Sciences (Pomona, CA) analyzed samples using The Testing Company LLC; a CLIA-certified California-based testing platform. Samples were extracted using the ThermoFisher Scientific MagMAX Viral/Pathogen Nucleic Acid Isolation Kit, and then analyzed using the Applied Biosystems$^{TM}$ 7500 Fast Dx Real-Time PCR Instrument using the CDC primers and probes against N1 and N2 genes: https://www.cdc.gov/coronavirus/2019-ncov/lab/rt-pcr-panel-primer-probes.html, and RNaseP as a positive control.

**Proof-of-Concept calibration of clinical samples:**

Similar to the first part of the harmonization study, this study is not a survey of test or method performance for limit of detection, precision, or repeatability; it is not a study of commutability. This study did not evaluate material homogeneity or stability. The primary goal of this study was to establish the reliability and reproducibility of inferring viral loads using calibrated samples run on different molecular testing platforms. Samples included 15 pools of 3 nasopharyngeal samples each, which had similar original Cq values for a total of 12 positive pools and 3 negative pools. The pools were shipped to 3 laboratories using biodesix (dPCR), biogazelle (qPCR), and LANL (qPCR), which used the same platforms described in the harmonization study. In addition, three secondary standards were shipped to each lab: Asuragen, LGC Clinical Diagnostics, and AcroMetrix (ThermoFisher Scientific), described in detail in **Supplementary Table 1**. Raw Cq values from the two qPCR platforms **(Supplementary Table 6A and 6B)** and counts from the dPCR platform **(Supplementary Table 6C)** were converted into IU measurements.

**Calibration analysis:**

To demonstrate the utility of commercially available harmonized SARS-CoV-2 calibrants, we conducted a “proof-of-concept” experiment using biobanked anterior nares clinical samples in viral transport media. We hypothesized that calibration of different PCR assays with harmonized reference materials will make quantitation of viral loads of clinical samples intercomparable. Three different labs measured 15 pooled clinical samples (ranging from clinical negative to high titer) and two calibration materials (Inactivated: AcroMetrix/ThermoFisher Scientific and Recombinant: AccuplexTM/LGC Clinical Diagnostics). We used the estimated IU/mL values for each reference from the harmonization study to assign the viral load estimates of the clinical samples using IU values. Samples were analyzed in three different labs using 5 different tests. We first compared calibrated results from both dPCR and qPCR on the same scale (IU) after calibration. We combined data from 5 experiments across the 3 labs. We analyzed the concordance of IU values extrapolated using 5 different PCR:Target combinations (dPCR: Biodesix:N1, Biodesix:N2, and qPCR: Biogazelle:E2:N, LANL:N1, and LANL: N2). We next compared IU/mL estimates of the clinical samples standardized using the AcroMetrix or LGC reference materials. The participating labs that provided raw data were not involved in the data analysis to minimize bias.

**Data Availability:**

Raw data and scripts are publicly available in this Github Repository: https://github.com/msalit/harmonization_dashboard. Processed data can be visualized using the Shiny app: https://msalit.shinyapps.io/RNAstudy/.
